# A Statistical Model for Quantifying the Needed Duration of Social Distancing for the COVID-19 Pandemic

**DOI:** 10.1101/2020.05.30.20117796

**Authors:** Nadav Rakocz, Boyang Fu, Eran Halperin, Sriram Sankararaman

## Abstract

Understanding the effectiveness of strategies such as social distancing is a central question in attempts to control the COVID-19 pandemic. A key unknown in social distancing strategies is the duration of time for which such strategies are needed. Answering this question requires an accurate model of the transmission trajectory. A challenge in fitting such a model is the limited COVID-19 case data available from a given location. To overcome this challenge, we propose fitting a model of SARS-CoV-2 transmission jointly across multiple locations. We apply the model to COVID-19 case data from Spain, UK, Germany, France, Denmark, and New York to estimate the distribution for the time needed for social distancing to end to range from May 2020 to July 2021 (95% credible interval), where the median date is October, 2020. Our method is not specific to COVID-19, and it can also be applied to future pandemics.

## 1 Introduction

Understanding the near-future implications of the COVID-19 pandemic is one of the most fundamental questions the scientific community is trying to answer in the past few months. While strategies such as social distancing have been widely employed to mitigate the impact of the pandemic on healthcare resources, the necessary timing, frequency, intensity, and effectiveness of these interventions is largely unknown. One of the key unknowns in these strategies is the duration of time for which social distancing needs to be imposed to flatten the pandemic curve. Answering this question requires an accurate model of the transmission trajectory of SARS-Cov-2.

Several recent studies [1, 2, 3, 4] have attempted to understand the transmission trajectory of SARS-CoV-2 using variants of compartmental models, such as the SEIR model [5, 6]. Prem et al. [1] fit an age-structured variant of the SEIR model to case data from Wuhan. They used this model to investigate the effect of lifting restrictions on returning to work and concluded that a premature and sudden lifting of interventions could lead to an early secondary peak. Li et al. [2] fit an SEIR model to SARS-CoV-2 case data across 375 cities in China during 10-23 January 2020. The model separately considered documented and undocumented infections. Further, they also integrated mobility data across cities. Using their model, they concluded that ≈ 86% of the cases were undocumented and these undocumented infections were the source of ≈ 80% of the documented cases. Kissler et al. [3] provide an elegant analysis using a variant of the SEIR model that takes into account various factors that modulate the transmission, including the effects of social distancing, seasonality, immunity, and cross-immunity, resulting in a highly detailed model that can predict, among other things, the time until social distancing is no longer required to flatten the curve. Using their model, they conclude that even under the assumption of full immunity as a response to infection, the time required for social distancing is at least 2022 assuming no vaccine or medication is found by then. Giordano et al. [4] consider a model with eight stages of infection: susceptible (S), infected (I), diagnosed (D), ailing (A), recognized (R), threatened (T), healed (H) and extinct (E). They apply their model to data from Italy to conclude that social distancing will need to be combined with testing and contact tracing to control the pandemic.

Unfortunately, even though the SEIR model is an established model, it is unclear to what extent the accuracy of the prediction of the time of social distancing is affected by the choice of the parameters. Further, the choice of parameters in these models in the context of SARS-Cov-2, has been a subject of debate within the scientific community. One of the key parameters that determine the transmission trajectory is the reproduction number, *R*_0_. Published values of *R*_0_ range from 1.4 to 7.23 [7, 8]. Kissler et al. chose to set peak *R*_0_ as ranging from 2.2 to 2.6, based on the fit of their model to historical data on related coronavirus (HCoV-OC43 and HCoV-HKU1) cases. This choice was made since at the time of publication, there was not enough SARS-COV-2 data to establish these parameters. However, currently, there is an opportunity to adjust the predictions based on SARS-COV-2 data as opposed to previous related viruses.

In this work, we fit a statistical model of transmission dynamics building upon the SEIR model. However, instead of fitting this model to previous strains of the SARS virus, we fit the model to data from current COVID-19 cases. A challenge with our approach arises from the limited case data available in a given location. Particularly, we demonstrate that the key epidemiological parameters that determine the end of social distancing (the reproduction number *R*_0_ and the average time spent in the infectious state *τ*) have large uncertainties associated with them which, in turn, lead to substantial uncertainties in estimates of the end of social distancing.

To obtain more precise parameter estimates, we formulate a hierarchical Bayesian model that allows the sharing of statistical strength across the location-specific models. Specifically, while each location is allowed to have its own values of the two parameters, these location-specific parameters are assumed to be drawn from a distribution centered around global parameter values. We estimate these global parameters using a marginal likelihood maximization framework. We then use these global parameter estimates, integrating over their uncertainty, to estimate the range of times till the end of social distancing in a new location. The resulting approach not only gives us point estimates (for parameters such as *R*_0_ and for the time to end social distancing) but also provides formal confidence intervals.

We apply our framework to COVID-19 cases from six locations (New York, Spain, Germany, France, Denmark, and the UK) to estimate global and location-specific parameter estimates. We show that these parameters provide a good fit to the data from each of the locations. Finally, we use the global parameter estimates to estimate that the time to end social distancing will be in October 2020 (assuming permanent immunity, no seasonality, and that social distancing reduces the effectiveness of transmission by 60%). We provide open-source software that can be applied to diverse locations to estimate transmission parameters and predict the required duration of social distancing. Although our analysis and motivation stems from the current COVID-19 pandemic, our method is general, and can be applied to other future pandemics.

## 2 Methods

### 2.1 The SEIR Model

We consider the extended SEIR model that have formed the basis of a number of recent studies of SARS-CoV-2 transmission dynamics[3]. This model partitions the population into susceptible, exposed but not yet infectious, infectious (mild), infectious (but not yet hospitalized), infectious (but not yet critical), hospitalized, critical (in the ICU), and removed. Given the state of the population at time *t, i*.*e*., the number of individuals in each of the partitions, the model describes the state of the population at the next time point by a set of ordinary differential equations which are governed by a number of parameters, such as the rate at which a susceptible individual is infected and rates at which an individual who is exposed becomes infectious, an infectious individual goes to the hospital, and so on (see Figure 1). Given the parameters and the state of the population at some initial time *t*_0_, this model allows us to compute the state of the population at subsequent times which, in turn, provides a trajectory of cases in the population.

**Figure 1:**
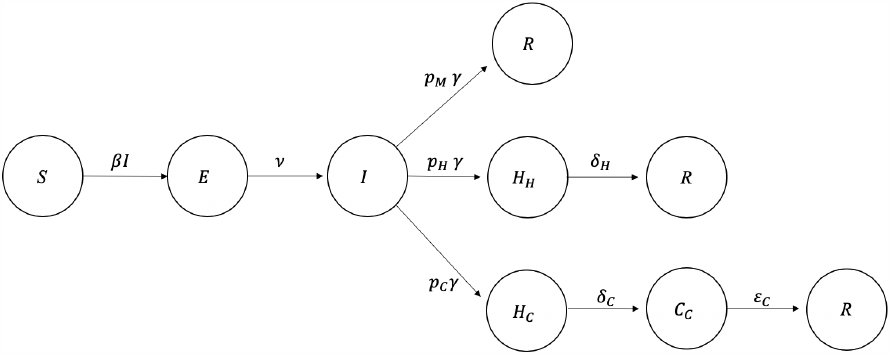
SEIR model schema. Each individual in the population begins at susceptible state *S*, and will enter into the exposure state *E* with transition rate *βI* in each time unit, where *β* = *R*_0_*γ*. In exposure state *E*, an individual will go into infectious state *I* with transition rate *ν*. Of all the people who arrive at state *I, p*_*M*_ of them will recover (state *R*), *p*_*H*_ will be hospitalized but will never reach critical care (state *H*_*H*_), and *p*_*C*_ will be hospitalized to later be in critical care (state *H*_*C*_). All transitions from the *I* state will occur with transition rate *γ*. People in *H*_*H*_ will enter into *R* with a transition rate *δ*_*H*_; people in *H*_*C*_ state will enter into critical state *C*_*C*_ with a transition rate *δ*_*C*_, and then enter into *R* state with a transition rate *∈*_*C*_. We set parameters *p*_*M*_ = 0.956, *p*_*H*_ = 0.0308, *p*_*C*_ = 0.0132, *ν* = 1*/*4.6, *δ*_*C*_ = 1*/*6, *δ*_*H*_ = 1*/*8, *E*_*C*_ = 1*/*10 as were estimated by Kissler *et al*. All states are normalized with respect to population size *N*.

Given the trajectory of SARS-CoV-2 cases from this model, a possible social distancing strategy involves imposing social distancing when the number of critical or hospitalized cases reaches the capacity of the health system and then relaxing social distancing when these numbers are sufficiently small. Depending on the transmission trajectory of SARS-CoV-2, social distancing may need to be imposed multiple times till a sufficiently large number of individuals in the population are immune (assuming that immunity to the virus is permanent). Social distancing is assumed to affect the transmission trajectory by changing the reproduction number *R*_0_.

The key parameters in this model that determine the time till the end of social distancing (*t*_*end*_) are the reproduction number (*R*_0_) and the average time during which an individual is infectious (*τ*). The parameter *τ* is related to the rate at which an individual transitions out of the infectious state typically used in the SEIR model (*γ*) as 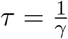.

### 2.2 A Bayesian hierarchical model for parameter estimation across multiple locations

Since it is unclear whether the parameters that fit HCoV-OC43 and HCoV-HKU1 are also applicable to SARS-Cov-2, we propose an alternate approach, in which we estimate the key parameter values by fitting the SEIR model to contemporary COVID-19 cases from specific locations. The challenge in such an approach is that the limited data in a given location leads to large uncertainty in the parameter estimates and is very sensitive to outliers.

Our approach to improving the precision of parameter estimates involves fitting SEIR models to all the locations jointly. One possible approach to do so involves setting the parameters to the same value across each location. However, this assumption is unlikely to be realistic. Instead, we endow each location-specific model with its own parameters but assume that the parameters are drawn from a distribution with global parameter values. The SEIR model has a number of parameters that control the transmission trajectory. Our model can jointly estimate all of these parameters. In our analysis, we fix all the parameters to values used in [3] but estimate the values of *R*_0_ and *τ*.

We assume that we have data on the observed number of COVID-19 cases from *K* locations: *{y*_*k*_(*t*)*}, t ∈ {*1, …, *T*_*k*_*}, k ∈ {*1, …, *K}*. Let *f* (*t*; (*R*_0_, *τ*)) denote the number of infections at time *t* predicted by the SEIR model with parameters (*R*_0_, *τ*). The parameters for each region are denoted 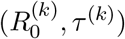 and the global parameters: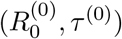.

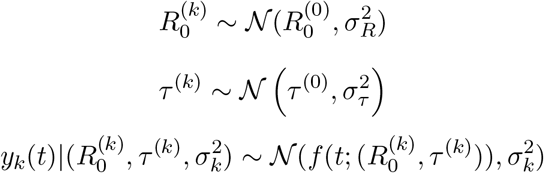

Each of the region-specific parameters is drawn from a normal distribution with a mean given by the global parameters. The observed cases in region *k* at time *t* are drawn from a normal distribution with mean given by the prediction from the SEIR model 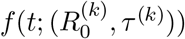 with a region-specific noise variance 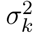. Further, we impose an uninformative prior on the noise variance: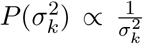. The parameter selection schema is shown as Figure 2

**Figure 2:**
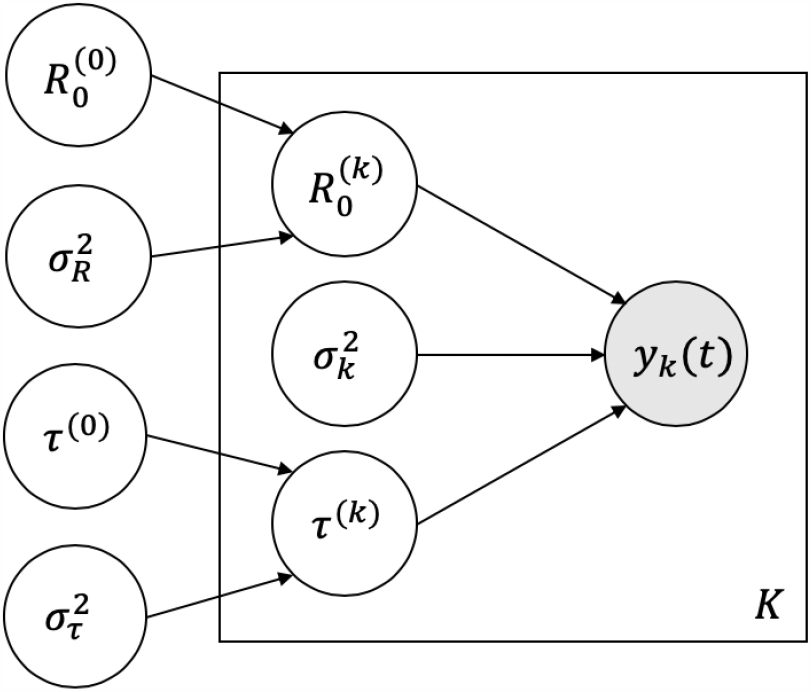
Parameter estimation diagram: We assume that the parameters 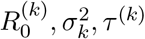 are drawn from a distribution which is defined by the parameters 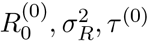, and 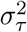 We then assume that the cumulative case number curve *y*_*k*_(*t*) is generated by the process defined by these parameters. We estimate the most likely values of 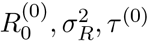, and 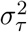 using maximum marginal likelihood approach.

We then have:

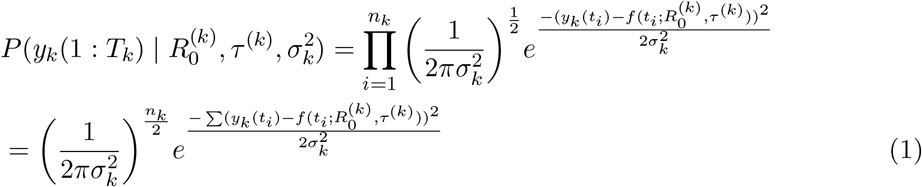

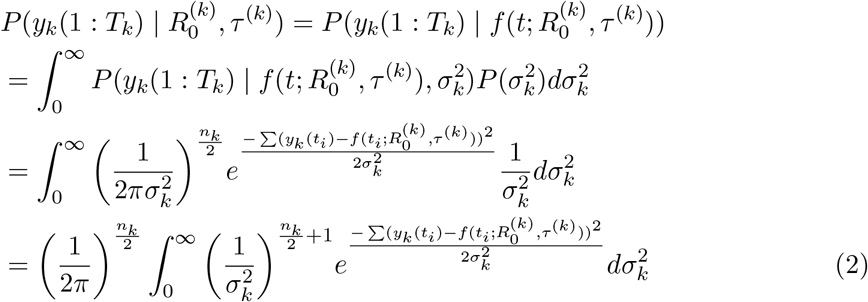

Using the fact that the integrand in Equation 2 is a Gamma function, we have:

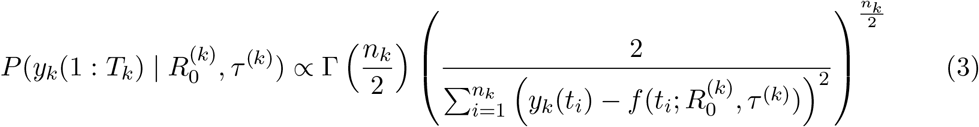

We then compute the maximum marginal likelihood estimates of the global parameters using a grid search:

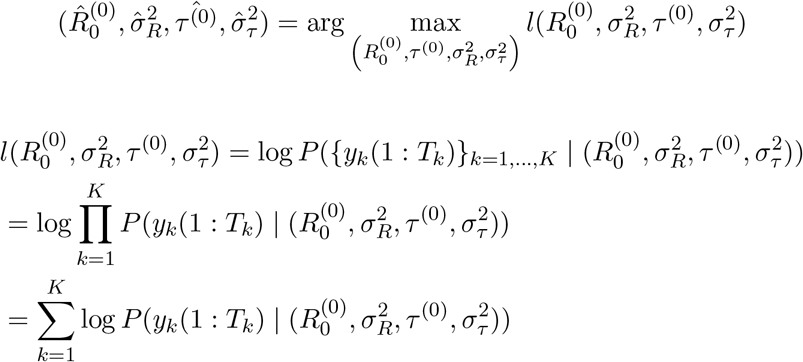

We evaluate each term in the log likelihood as:

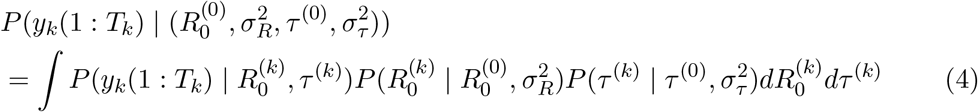

The integral in Equation 4 does not have an analytical solution so we evaluate the integral numerically over a grid of values for 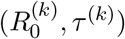.

The grid search of the parameters in the likelihood searches for values of 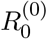 between 1 and 8, *τ* ^(0)^ between 2 to 55, *σ*_*R*_ from 1 to 8, and *σ*_*τ*_ from 1 to 30.

### 2.3 Application to predict the end of social distancing

We estimate *t*_*end*_, the time when social distancing can be ended, in the following way. First, using a maximum marginal likelihood approach, we find the most likely parameters 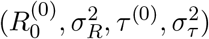. Then, we sample *R*_0_, *τ* from the distribution 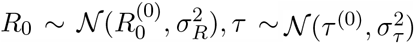 and for each such sample we compute the estimated value of *t*_*end*_ as follows. We follow the parameter choices used in [3]: assuming that immunity to SARS-CoV-2 is permanent (which provides the minimum time of social distancing), that social distancing is imposed when the number of cases exceeds 35 per 10, 000 individuals and is relaxed when the number of cases drops below 5 per 10, 000 individuals (these thresholds were chosen so that the number of hospital cases is below the capacity in the United States), and that each period of social distancing reduces *R*_0_ by 60%. We then simulate the SEIR scenario based on the above parameters, including *R*_0_ and *τ*. This result in a distribution of values *t*_*end*_. Additionally, we performed a sensitivity analysis where we demonstrate the effect of the choice of each of the above parameters.

## 3 Results

### 3.1 Estimates of *t*_*end*_ from region-specific parameter estimates

We consider COVID-19 data[9, 10] from six locations: UK, Spain, Germany, France, Denmark, and New York. Since our goal is to estimate the parameters (*R*_0_ and *τ*) in the period when no social distancing was imposed, we restricted our analysis to the dates prior to when social distancing was imposed in each of these regions.

Figure 3 shows the parameter estimates when we fit a SEIR model to each of the six regions. While each of the models appears to fit the data in each of the regions, there is considerable uncertainty in the parameter estimates (see Table 1 for 95% CI). We note that the uncertainty in the key epidemiological parameters that determine the end of social distancing (the reproduction number *R*_0_ and the average time spent in the infectious state *τ*) leads to substantial uncertainties in estimates of *t*_*end*_: the time till the end of social

**Table 1:** The maximum-likelihood estimates for R0 and *τ* for every region distancing (Figure 4 and Figure 5).

| Region | $R_0$ | $\tau$ |
| --- | --- | --- |
|  | [95% confidence interval] | [95% confidence interval] |
| UK | 7.6 [6.6,8.0] | 27 [26,31] |
| New York | 7.9 [4.5,8.0] | 4 [2,4] |
| Spain | 7.6 [5.6,8.0] | 11 [10,11] |
| France | 8.0 [7.1,8.0] | 35 [29,35] |
| Germany | 8.0 [6.9,8.0] | 28 [23,28] |
| Denmark | 8.0 [1.0,8.0] | 5 [2,5] |

**Table 2:** The date of the end of social distancing for different sets of parameters that fit the data (as shown in Figures 3 and 4)

| Region used for<br>parameter estimation | $R_0$ | $\tau$ | Estimated end of<br>social distancing |
| --- | --- | --- | --- |
| United Kingdom | 1.1 | 10 | Apr, 2020 |
| New York | 7.9 | 4 | May, 2020 |
| Denmark | 8.0 | 5 | Jun, 2020 |
| Denmark | 5.4 | 5 | Jul, 2020 |
| Spain | 7.6 | 11 | Aug, 2020 |
| New York | 4.2 | 3 | Sep, 2020 |
| Denmark | 1.6 | 2 | Nov, 2020 |
| United Kingdom | 7.6 | 27 | Jan, 2021 |
| Germany | 7.4 | 28 | Feb, 2021 |
| United Kingdom | 4.6 | 20 | Apr, 2021 |
| France | 7.6 | 34 | Apr, 2021 |
| Spain | 3.9 | 7 | Aug, 2021 |
| Germany | 4.0 | 16 | Aug, 2022 |
| Spain | 3.4 | 7 | Sep, 2022 |
| France | 4.2 | 26 | Feb, 2023 |
| Germany | 3.0 | 19 | Feb, 2024 |
| France | 1.8 | 19 | Mar, 2024 |

**Figure 3:**
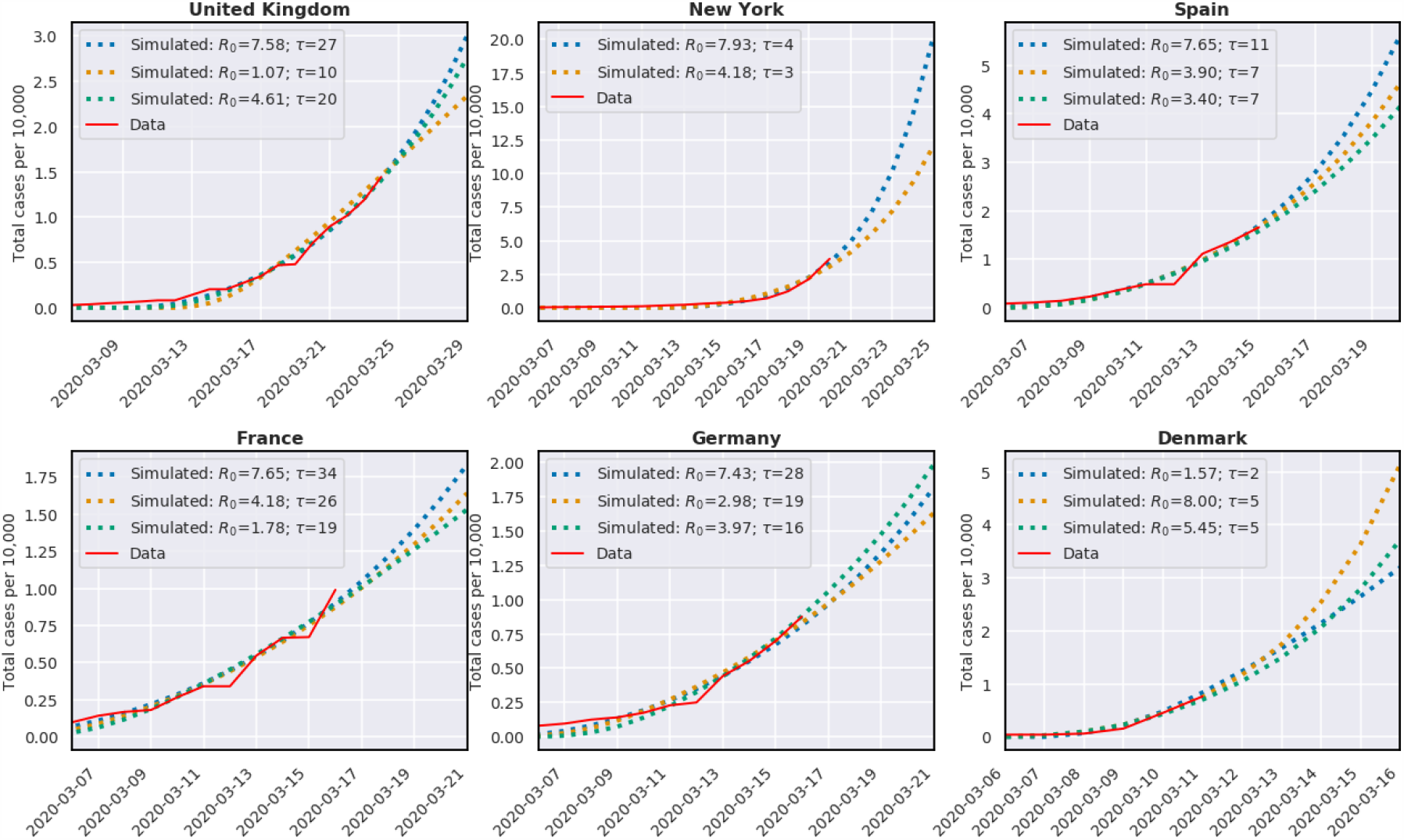
Comparison of the observed trajectory of the number of cases in United kingdom, New York, Spain, France, Germany, and Denmark (prior to the date where social distancing was imposed). We provide fits based on region-specific parameters (we choose sets of parameters that all lie within the 95% confidence set). The different sets of parameters diverge significantly in the subsequent dates showing the under-determination of this model.

**Figure 4:**
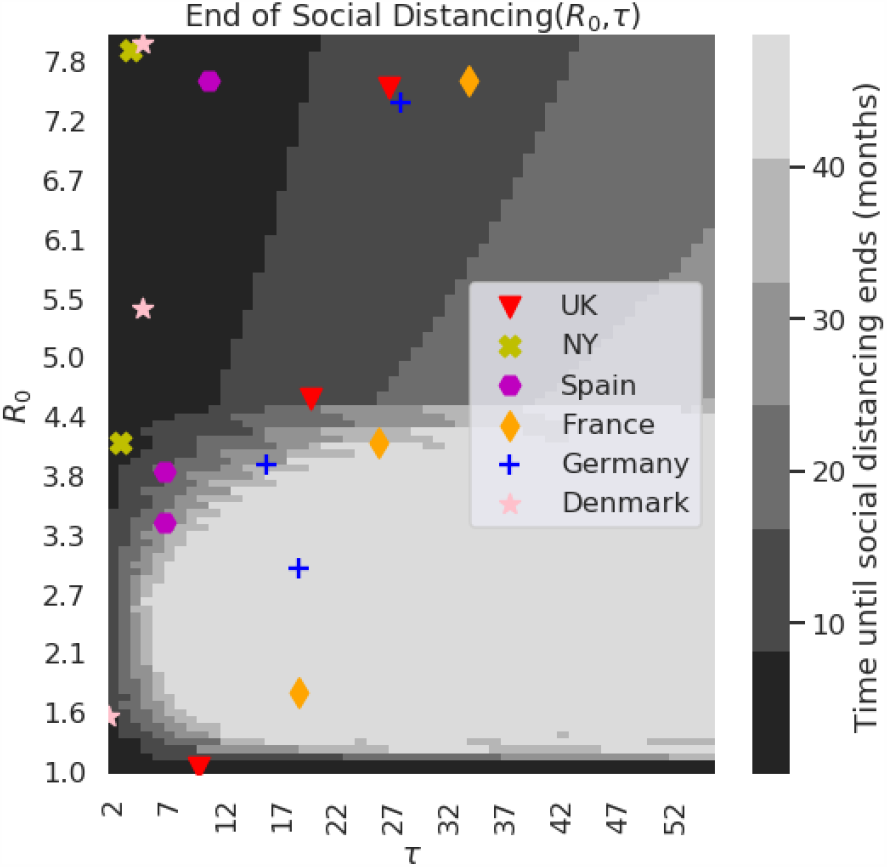
The time until social distancing ends (in months) based on the SEIR model, using different *R*_0_ and *τ* values. For each of the regions (Spain, United Kingdom, New York, France, Germany, and Denmark) we also marked the parameters that provided a good fit as shown in Figure 3.

**Figure 5:**
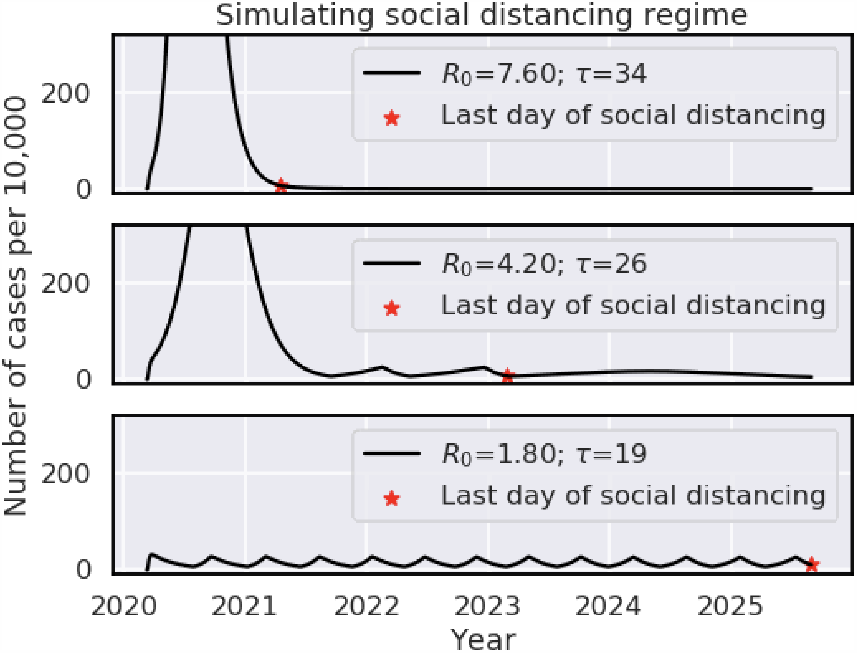
Simulating the number of cases under the social distancing regime where social distancing is turned on when the number of cases exceeds 35 per 10, 000 and is turned off when it drops below 5 per 10, 000. We show 3 different sets of parameters matching data taken from France as seen in Table 2.

### 3.2 Estimates of *t*_*end*_ using a Bayesian framework

Due to the large uncertainty in the parameters estimated in each of the locations separately, we fit our model jointly in all locations using a Bayesian framework (see Methods). The Bayesian framework assumes a prior distribution (normal) on the parameters 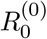 and *τ*^0^, and it estimates the posterior probability based on the data obtained in each of the countries. The estimated global parameters of the model are 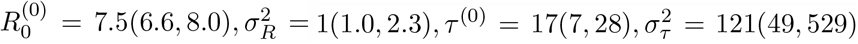. We observe that our parameter estimates provide an adequate fit to the data in each of the locations (Figure 6). We then sample the parameters from the most likely distribution of the parameters (*R*_0_, *τ*) and for each set of parameters we simulate the pandemic scenario, while taking into account that social distancing reduces *R*_0_ by 60%, and under the assumption that immunity is fixed for life once exposed. The latter assumption is a best-case scenario, i.e., if this assumption is relaxed then the time to social distancing is expected to increase. Furthermore, we assume no seasonality, and again, this results in a lower bound on the time for social distancing. However, since we do not have any strong evidence for specific effects of seasonality, or specific information about the duration of immunity, we chose to focus on this lower bound scenario. Under this scenario, our analysis provides a distribution of possible values for *t*_*end*_ (Figure 7). The mode of the distribution is in September 2020, the median is in October 2020 and the variance is 16 months. Based on these results, we obtain a more optimistic view of the time for the end of social distancing compared to previous analysis [3].

**Figure 6:**
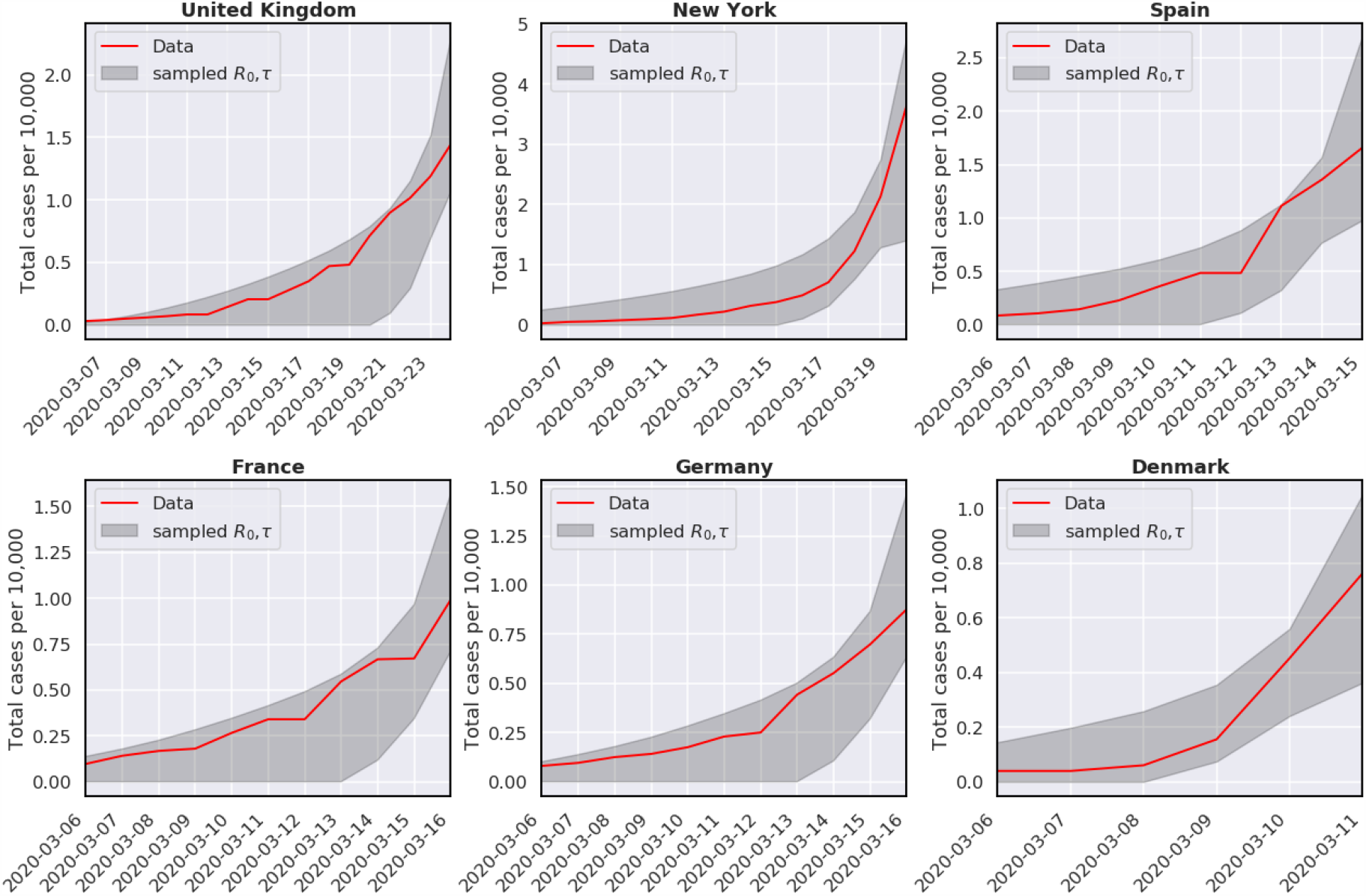
The range of trajectories for the number of cases predicted using samples from distribution implied by the global parameters estimated on all regions.

**Figure 7:**
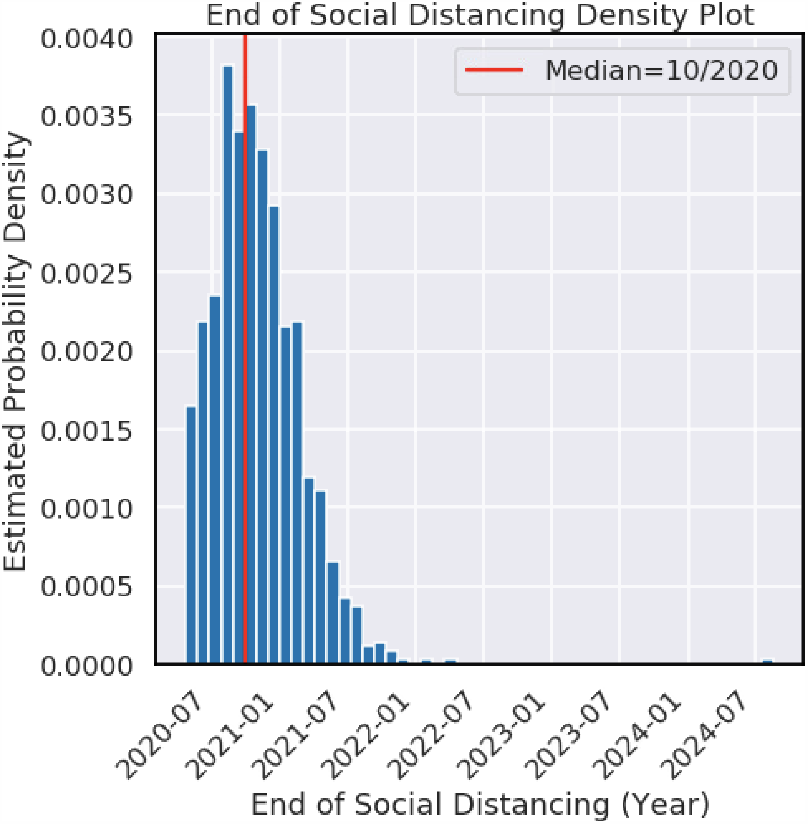
The distribution for the time until social distancing will end implied by the global parameters 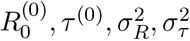. The median is October 2020, the mode is September 2020, the variance is 16 months, and the 95% credible interval ranges from May 2020 to July 2021.

### 3.3 Sensitivity analysis

We first wanted to check how our estimates of *t*_*end*_ are affected by the choice of the specific regions. Out of the six regions (United Kingdom, New York, Spain, France, Germany, and Denmark) we iteratively chose four regions and estimated the global parameters 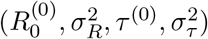. For each such set of parameters, we estimated the median of the time for social distancing by sampling 1000 samples from the distribution implied by these parameters, resulting in 1000 estimates of the time in which the social distancing will end. We observe that the median *t*_*end*_ is not greatly affected by the choice of the regions, and particularly the medians typically range from September 2020 to April 2021 (Figure 8 (a)).

**Figure 8:**
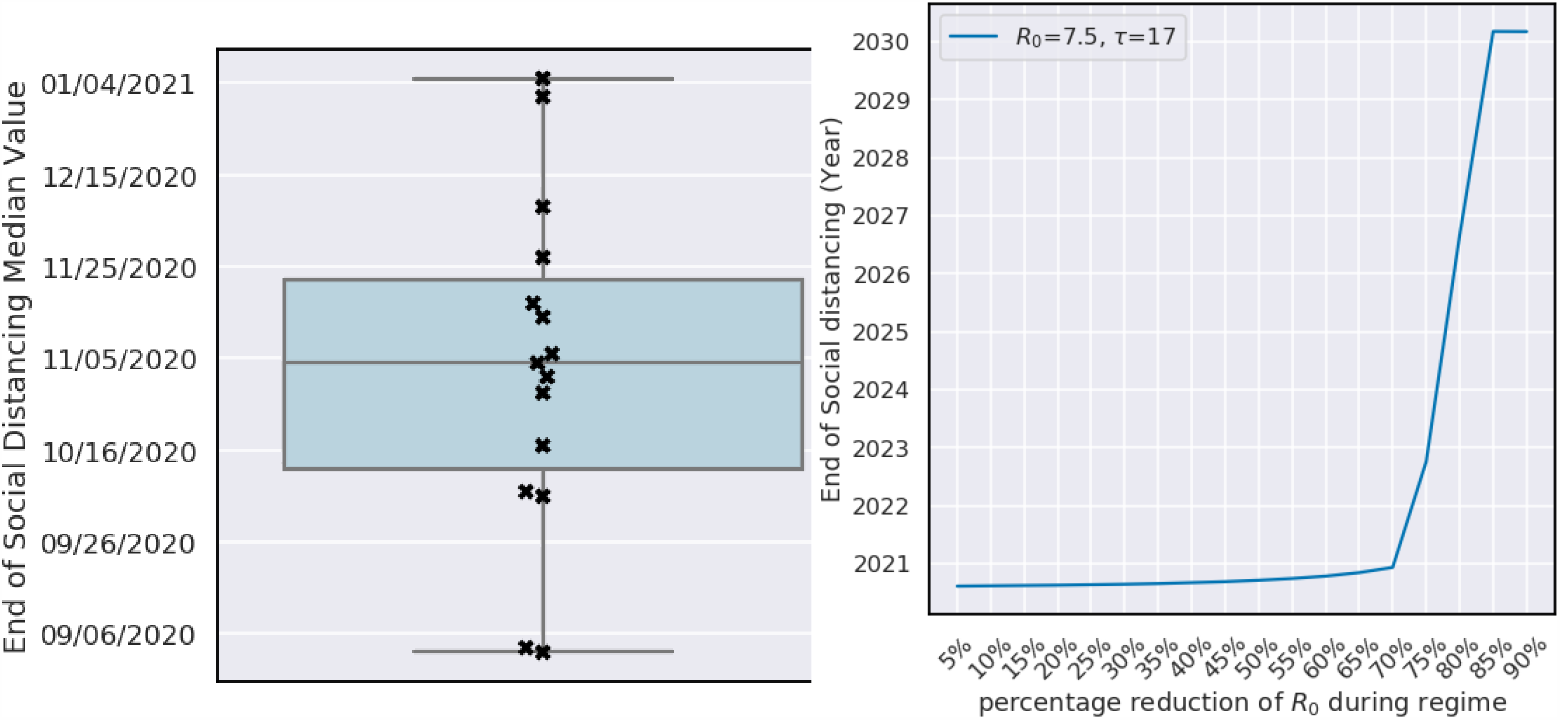
(a) Different times until social distancing will end based on different choices of the regions. Each sample was generated by choosing four regions out of the six (UK, France, Spain, Germany, New York, and Denmark), estimating their global parameters, and then measuring the median for the time social distancing will end implied by these parameters. (b) End of social distancing regime as a function of the percentage of *R*_0_ during the regime.

We next wanted to examine the effect of the decrease in *R*_0_ as a result of social distancing on our estimates. We therefore fixed the values of *R*_0_ and *τ* to the maximum marginal likelihood estimates (*R*_0_ = 7.5 and *τ* = 17), and varied the effect of social distancing on *R*_0_. Interestingly, this results in a phase transition behavior where the time for social distancing will end within the next year if social distancing has a moderate effect (*i*.*e*., it reduces *R*_0_ by less than 60%), or it will end within many years if social distancing has a large effect (Figure 8 (b)).

## 4 Discussion

In this work, we fit a statistical model of transmission dynamics based on the SEIR model to data from COVID-19 cases from multiple locations. Our approach uses a Bayesian framework, resulting in a distribution of end dates for social distancing, as opposed to a specific end time, incorporating the uncertainty in the parameter choices of the model. This uncertainty is inherent to the SARS-Cov-2 pandemic, as can be viewed by the fact that *R*_0_ has been ranging in the literature from 1.4 to 7.23 [7, 8]. We show that our approach provides a good fit for the COVID-19 cases in these locations. Our approach demonstrates that the end of social distancing will be around October 2020, under mild assumptions.

It is important to note that the assumptions made by our analysis provide a lower bound on the time for social distancing. Particularly, we assume no seasonality; if COVID-19 is seasonal, we expect greater spread to appear in winter relative to summer (as has been observed for influenza). However, it is not clear whether COVID-19 is seasonal, and if so, to what extent, and we therefore leave this aspect for future analysis, once more data will be available. Similarly, it is currently unclear whether one acquires permanent immunity or for a short duration after getting exposed to the disease. Thus, given the lack of information about immunity, we chose to make the best-case scenario assumption in which immunity is acquired for life. Other assumptions may prolong the effects of social distancing.

Critically, other interventions such as the introduction of a vaccine, the introduction of effective medications, or the introduction of a larger number of clinical care resources such as ventilators, will change the scenarios provided in this analysis. Specifically, the introduction of a vaccine or effective medications will likely alter the parameters *R*_0_ and *τ*, and would therefore result in a shorter time for social distancing. Additional resources such as ventilators will result in different thresholds set for the application of social distancing, as social distancing will only be required when there is a risk for a surge that collapses the health systems. In that case, again, the end of social distancing is expected to arrive sooner.

The above limitations need to be taken into account when interpreting our analysis. However, we note that as new data on immunity, seasonality, medications, and vaccines becomes available, these can easily be incorporated into our framework, and a revised analysis can be performed. We provide freely available code that allows for such an analysis by researchers in the community (see appendix).

Finally, we would like to point out that the issue of sensitivity of the model to the parameter choices is not specific to the SEIR model. Specifically, in our hands we have observed a similar phenomenon for other models as well (data not shown). The limited availability of data limits the certainty with which parameters of the model can be identified. Thus, it is critical that estimates from the application of statistical models to such data be accompanied by formal measures of uncertainty. We believe that the statements resulting from our analysis should also be taken in the context of the specific locations we analyzed and the specific model that we used. Possibly, other models or other locations may provide different estimates.

## Data Availability

Public COVID-19 data used to generate the experiments presented in this manuscript were acquired from the Johns Hopkins University and New York Times COVID-19 GitHub repositories.

https://github.com/CSSEGISandData/COVID-19

https://github.com/nytimes/covid-19-data

## A SEIR Model

### A.1 Ordinary differential equations

The SEIR model described by Kissler *et al*. is defined by the following set of ordinary differential equations:

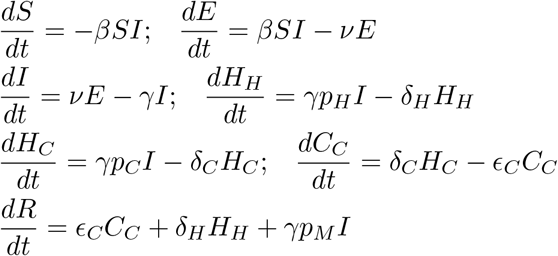

### B Code availability

The code used to generate all figures and experiments in this paper can be found here: https://github.com/doubleBlindGit/COVID19_SocialDistance

## Notes

### Competing Interest Statement

The authors have declared no competing interest.

### Funding Statement

no external funding was received

### Author Declarations

No IRB required

